# Estimating effects of physical distancing on the COVID-19 pandemic using an urban mobility index

**DOI:** 10.1101/2020.04.05.20054288

**Authors:** Jean-Paul R. Soucy, Shelby L. Sturrock, Isha Berry, Duncan J. Westwood, Nick Daneman, Derek R. MacFadden, Kevin A. Brown

**Author notes:** Correspondence to: Jean-Paul R. Soucy Division of Epidemiology, Dalla Lana School of Public Health, University of Toronto 155 College St Room 500, Toronto, ON M5T 3M7 Canada (1).

## Abstract

**Background:** Governments have implemented population-wide physical distancing measures to control COVID-19, but metrics evaluating their effectiveness are not readily available.

**Methods:** We used a publicly available mobility index from a popular transit application to evaluate the effect of physical distancing on infection growth rates and reproductive numbers in 40 jurisdictions between March 23 and April 12, 2020.

**Findings:** A 10% decrease in mobility was associated with a 14.6% decrease (exp(β) = 0·854; 95% credible interval: 0·835, 0·873) in the average daily growth rate and a −0·061 (95% CI: −0·071, −0·052) change in the instantaneous reproductive number two weeks later.

**Interpretation:** Our analysis demonstrates that decreases in urban mobility were predictive of declines in epidemic growth. Mobility metrics offer an appealing method to calibrate population-level physical distancing policy and implementation, especially as jurisdictions relax restrictions and consider alternative physical distancing strategies.

**Funding:** No external funding was received for this study.

**Research in Context:** *Evidence before this study:* Widespread physical distancing interventions implemented in response to the COVID-19 pandemic led to sharp declines in global mobility throughout March 2020. Real-time metrics to evaluate the effects of these measures on future case growth rates will be useful for calibrating further interventions, especially as jurisdictions begin to relax restrictions. We searched PubMed on May 22, 2020 for studies reporting the use of aggregated mobility data to measure the effects of physical distancing on COVID-19 cases, using the keywords “COVID-19”, “2019-nCoV”, or “SARS-CoV-2” in combination with “mobility”, “movement”, “phone”, “Google”, or “Apple”. We scanned 252 published studies and found one that used mobility data to estimate the effects of physical distancing. This study evaluated temporal trends in reported cases in four U.S. metropolitan areas using a metric measuring the percentage of cell phone users leaving their homes. Many published papers examined how national and international travel predicted the spatial distribution of cases (particularly outflow from Wuhan, China), but very little has been published on metrics that could be used as prospective, proximal indicators of future case growth. We also identified a series of reports released by the Imperial College COVID-19 Response Team and several manuscripts deposited on preprint servers such as medRxiv addressing this topic, demonstrating this is an active area of research.

*Added value of this study:* We demonstrate that changes in a publicly available urban mobility index reported in over 40 global cities were associated with COVID-19 case growth rates and estimated reproductive numbers two to three weeks later. These cities, spread over 5 continents, include many regional epicenters of COVID-19 outbreaks. This is one of only a few studies using a mobility metric applicable to future growth rates that is both publicly available and international in scope.

*Implications of all the available evidence:* Restrictions on human mobility have proved effective for controlling COVID-19 in China and the rest of the world. However, such drastic public health measures cannot be sustained indefinitely and are currently being relaxed in many jurisdictions. Publicly available mobility metrics offer a method of estimating the effects of changes in mobility before they are reflected in the trajectory of COVID-19 case growth rates and thus merit further evaluation.

## Introduction

Policies limiting contact between individuals outside of households, via school closure, voluntary telecommuting, and shelter-at-home orders, have been implemented throughout most of the world to reduce the transmission of COVID-19. Physical distancing (previously termed social distancing) policies have helped control previous epidemics of respiratory infections^1,2^ and played a significant role in reducing COVID-19 transmission in China.^3,4^ Many regions have adopted physical distancing measures, incrementally increasing restrictions and enforcement overtime. Gradual relaxation of these measures has now begun in many jurisdictions, but some version of physicial distancing is likely to continue into the near future in order to control the pandemic.^5^ A proximal indicator of future infection rates (at a known time lag) is urgently needed to guide the future implementation and modification of physical distancing and other non-pharmaceutical interventions. In this analysis, we demonstrate that a mobility index based on regular users of a web-based transit application can capture the effect of physical distancing on the reproductive number and growth rate of COVID-19 in 40 states and countries spanning 5 continents.

We used a daily city-level mobility index to (a) measure adherence to large-scale movement restrictions, and (b) predict the COVID-19 growth rate and instantaneous reproductive number at the national and sub-national level. The mobility index was provided by a public transit application (app) and uses the number of trips planned in the app to estimate the percentage of each city that is commuting relative to an internal reference from a recent usage period. The index is available from March 2^nd^ to present and includes all 41 cities where the app operates. Importantly, outbreaks in major urban centers (like those in the dataset) represent a large proportion of total COVID-19 cases at national and sub-national (regional) levels^6^. As a result, reduced mobility in these cities should have a significant impact on infection growth rates at larger geographies. Further, changes in the city-level mobility index are related to physical distancing interventions, many of which were implemented at the national or sub-national level^7^; thus, reduced mobility in major urban centers should serve as a reasonable proxy for larger-scale behavior change.

## Methods

### Mobility index

The Citymapper Mobility Index (CMI; https://citvmapper.com/cmi) includes data on 41 cities in 23 countries. CMI measures the relative frequency of trips planned within the application in 41 cities across the Americas, Europe, Australia and Asia, compared to an internal reference at the beginning of 2020 (or at the end of 2019, in the case of Singapore and Hong Kong). The CMI is available from March 2^nd^ to present.

To validate our use of CMI as a measurement for adherence to physical distancing measures, we plotted the CMI in each city before and after the first announcement of major national or sub-national physical distancing interventions, namely gathering restrictions and/or mandatory closures (figure 1, table S1). Unlike a declaration of emergency, these measures have clear and consistent implications across regions. The announcement of physical distancing measures generally coincided with steep drops in mobility throughout the month of March, although in many cases mobility was declining to some extent prior to these announcements.

**Figure 1.**
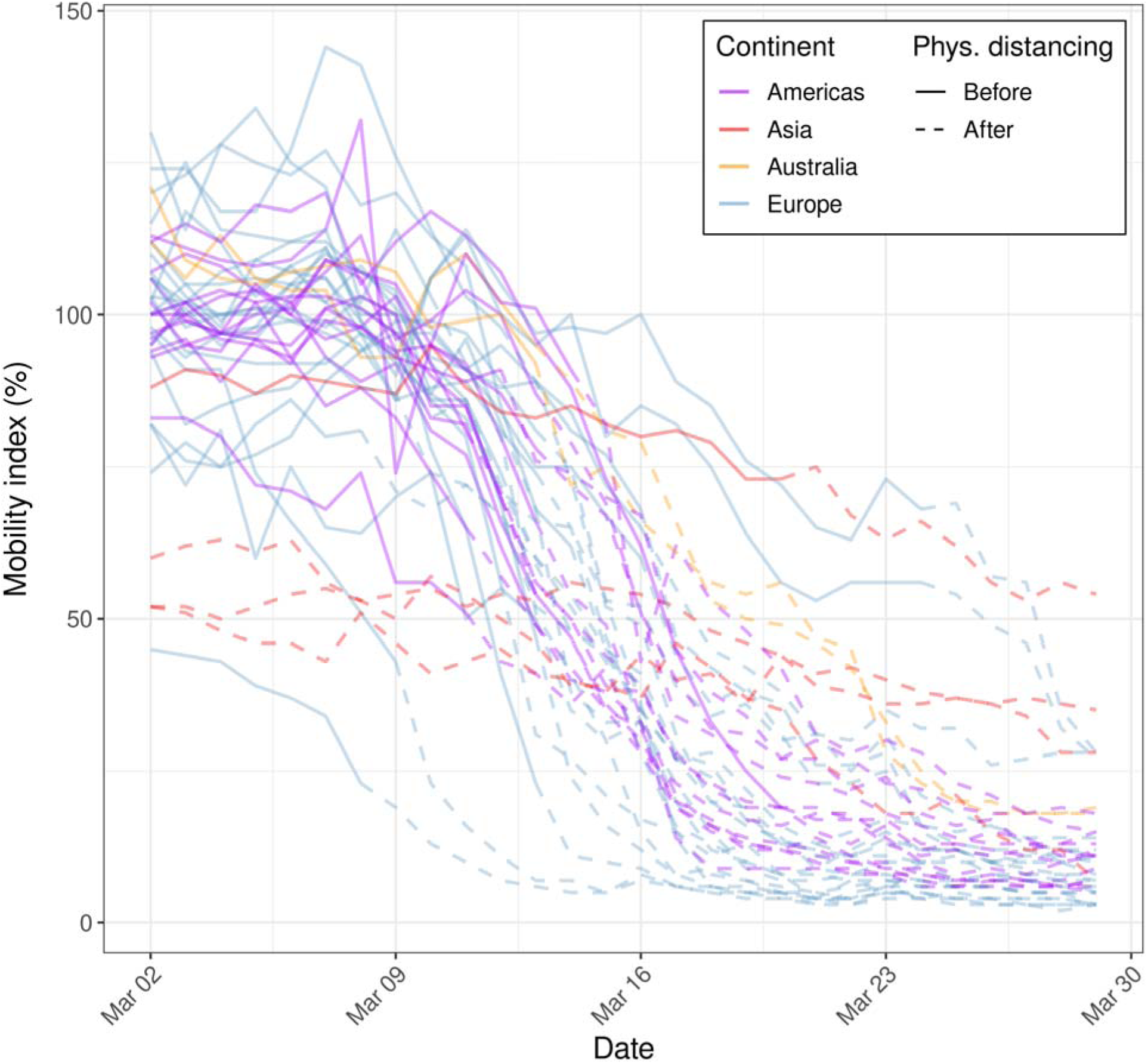
Mobility index in 41 cities over a four-week period in March 2020, before (solid lines) and after (dashed lines) the first major state- or country-level physical distancing intervention was announced.

The CMI dataset contained two French cities (Paris and Lyon), but region-level cumulative case counts were not available for France for the entire study period. In subsequent analyses, we calculated a populated weighted CMI for all of France based on the two cities· metropolitan area populations.

### COVID-19 case growth rates

We obtained national and sub-national (where available) cumulative case time series for countries represented in the Citymapper dataset (table S2). In all, we obtained 40 regional- and national-level cumulative case time series. We calculated daily growth rates for each region (presented as a percentage) by dividing the number of new cases reported in a given day by the cumulative number of cases as of the previous day. In our models, we used three weeks of case data, beginning with the week of March 23, 2020 and concluding with the week of April 6, 2020.

We used multilevel linear regression with random intercepts for locations nested within countries (to account for the non-independence of observations from the same location and for the clustering of sub-national units within a country, respectively) to estimate the association between the logarithm of the mean daily growth rate and mean CMI in prior weeks. All models were fit using Stan^8^ via the rstanarm package (version 2·19·3)^9^ in R (version 3·6·3).^10^ For all models, we used the default settings and default scaled, weakly informative priors for all parameters.

Based on the known lag between infection and symptom onset of 5·4 days^11^, plus an estimated lag between symptom onset and public reporting of 5 to 15 days (e.g., 6·4 days in Singapore up to March 17^12^), we used a two-week lag as our primary analysis, with a three-week lag as a sensitivity analysis. Most cities implemented their first major physical distancing policies throughout the second week of March (March 9-March 15). Thus, we compared average weekly mobility (for the weeks of March 9, March 16, and March 23) to the growth rate of cases two weeks later (the weeks of March 23, March 30, and April 6, respectively), both aggregated by week, for 40 states and countries. For most countries in the dataset, this period represents the growth and/or early peak phases of the epidemic.^6^

To adjust for epidemic timing, we ran an additional model including days since the 100^th^ case as a continuous covariate (excluding the Principality of Monaco, which had fewer than 100 cases by the end of the study period). However, we note that epidemic timing is closely related to the implementation of physical distancing and reductions in mobility. For example, Italy, which had an earlier epidemic than other European countries, had a substantially lower CMI in early March than other European countries in the dataset.

As a sensitivity analysis, we re-ran the model using median daily growth rate as the outcome. This method is less influenced by outliers, such as the one caused by a change in case definition in Quebec, Canada on March 23.

### COVID-19 instantaneous reproductive number

The instantaneous reproductive number is a quantity signifying the average number of secondary infections a person infected at time *t* would be expected to generate given that conditions remain unchanged.^13^ We estimated the instantaneous reproductive for the time intervals corresponding to the weeks of March 23, March 30, and April 6 using the EpiEstim package (version 2·2-1)^14^ in R and daily incidence from March 8 to April 12. We employed the parametric serial interval method^15^ using parameters from Du and colleagues^16^ (mean = 3·96 days, SD = 4·75 days).

We used multilevel linear regression with random intercepts for locations nested within countries to estimate the association between the calculated instantaneous reproductive number and mean CMI in prior weeks. We used a two-week lag as our primary analysis with a three-week lag as a sensitivity analysis. To adjust for time since epidemic start, we ran an additional model including days since the 100^th^ case as a continuous covariate (excluding the Principality of Monaco, which had fewer than 100 cases at the end of the study period).

### Imported cases

Data on imported versus locally acquired cases were not available for most locations, so we were unable to adjust for this factor during the analysis. As a sensitivity analysis, we re-ran the models by excluding the first week (March 23) of case data. Cases reported during this first week are much more likely to represent imported cases than cases reported during later weeks, since travel restrictions were implemented during the third week of March in many countries.^17^

## Results

### Mobility index

Nearly all cities experienced substantial reductions in mobility during the month of March (March 2: mean = 97·6%, SD = 19·0; March 29: mean = 12·7%, SD = 10·4) (figure 1). Decreases were less pronounced in Hong Kong and Seoul, where mobility was already substantially reduced at the beginning of the month. Cities within Europe, Australia, and the Americas showed strikingly similar patterns in mobility reduction that corresponded to the dates of national or sub-national physical distancing mandates, including restrictions on public gatherings or mandatory closures (table S1). Increasingly restrictive physical distancing policies were adopted in an incremental fashion following the index date and thus continued declines in mobility were expected through the remainder of the month. Messaging from public health authorities and news media likely contributed to changes in behavior prior to the index date.

### COVID-19 case growth rates

The mean mobility index was associated with the logarithm of the growth rate of cumulative cases 2 weeks later (figure 2). A 10% lower mean mobility index was associated with a 14.6% lower mean daily growth rate two weeks later (exp(β) = 0·854; 95% credible interval (CI): 0·835, 0·873) (table 1). When the model was adjusted for days since the 100^th^ case (a measure of epidemic timing), the association was attenuated (exp(β) = 0·955; 95% CI: 0·926, 0·985). However, epidemic timing and mobility are closely related.

**Figure 2.**
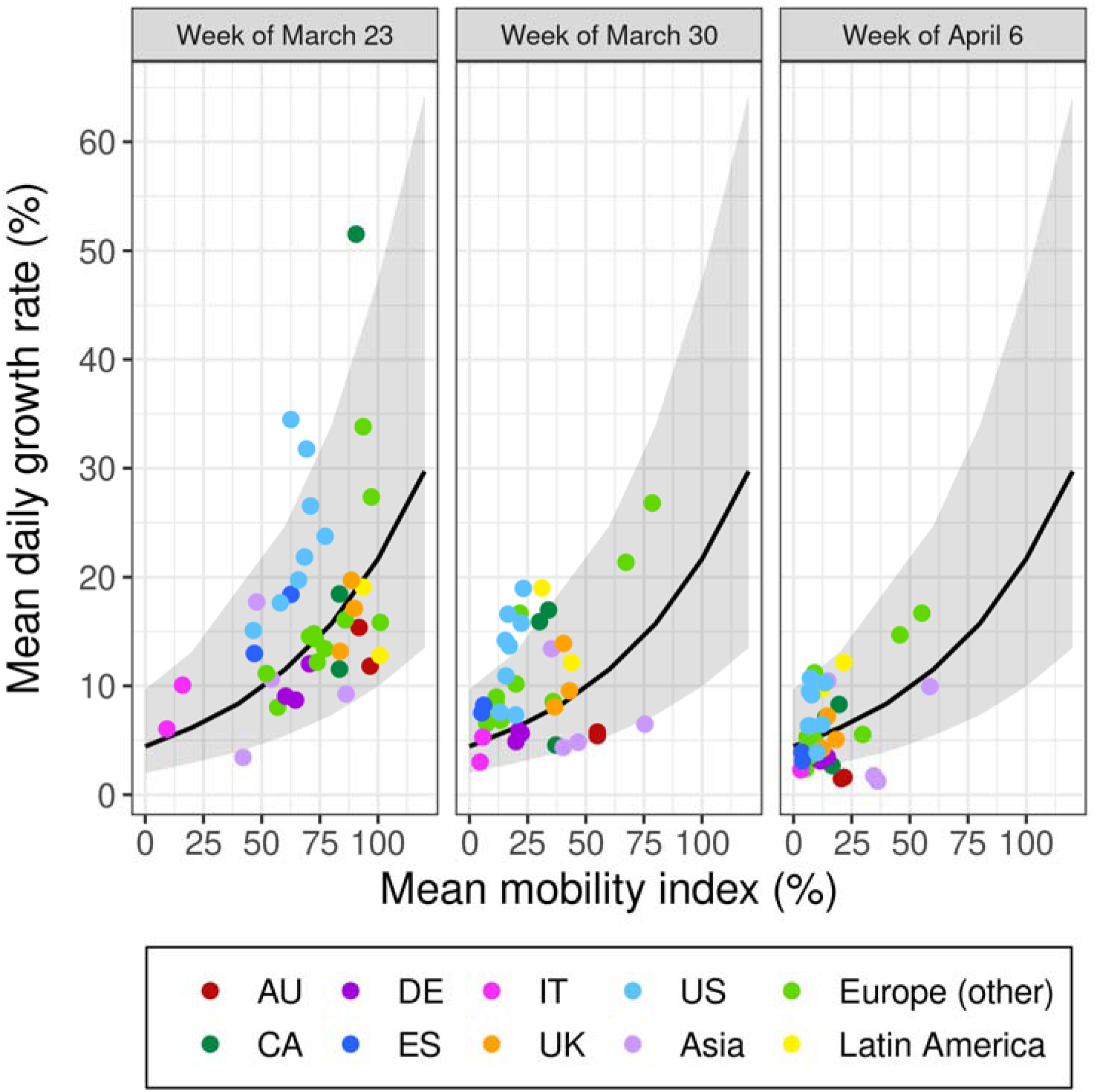
The association between mean daily growth rate (aggregated by week) and mean mobility index two weeks prior for 40 states and countries. Each pair of weeks (labeled according to the week growth rate was measured) is plotted separately for a model fit on all three weeks of data. The 95% prediction interval is shown in gray. AU = Australia; CA = Canada; DE = Germany; ES = Spain; IT = Italy; UK = United Kingdom; US = United States.

**Table 1.** Model coefficients (with 95% credible intervals) for the association between a 10% decrease in the mobility index and the mean daily growth rate and instantaneous reproductive number (both aggregated by week) assuming a lag of two or three weeks. Model coefficients are presented with and without adjustment for days since 100^th^ case. Models include three weeks of outcome data (March 23-April 12) for 40 countries and states (39 in the adjusted models). CI = credible interval.

| **Mean daily growth rate (%) ($\exp(\beta)$)** |  |  |  |
| --- | --- | --- | --- |
| **Mobility index** | **Unadjusted (95% CI)** | **Adjusted (95% CI)<sup>a</sup>** |  |
|  | **Mobility<sup>b</sup>** | **Mobility<sup>b</sup>** | **Days since 100 cases<sup>c</sup>** |
| 2-week lag | 0.854 (0.835, 0.873) | 0.955 (0.926, 0.985) | 0.558 (0.490, 0.639) |
| 3-week lag | 0.865 (0.851, 0.880) | 0.941 (0.913, 0.969) | 0.620 (0.534, 0.720) |
| **Instantaneous reproductive number ($\beta$)** |  |  |  |
| **Mobility index** | **Unadjusted (95% CI)** | **Adjusted (95% CI)<sup>a</sup>** |  |
|  | **Mobility<sup>b</sup>** | **Mobility<sup>b</sup>** | **Days since 100 cases<sup>c</sup>** |
| 2-week lag | -0.061 (-0.071, -0.052) | -0.029 (-0.044, -0.014) | -0.169 (-0.233, -0.108) |
| 3-week lag | -0.052 (-0.060, -0.044) | -0.020 (-0.035, -0.005) | -0.183 (-0.261, -0.108) |

<sup>a</sup> Excludes the Principality of Monaco.
<sup>b</sup> Coefficient for a 10% decrease in the mobility index.
<sup>c</sup> Coefficient for a 10-day increase since the 100<sup>th</sup> reported case.

Our findings were robust to estimation of median daily growth rate (exp(β) = 0·862; 95% CI: 0·844, 0·880), which reduces the influence of outliers. Since reporting delay is unknown and variable across geographies, we also ran a model using a three-week lag for mobility, which produced a similar strength of association (table 1). We were unable to exclude imported cases, which were likely most prevalent during the first week of case data (March 23). Re-running models to exclude this week produced relatively comparable results (table S3).

### COVID-19 instantaneous reproductive number

The mobility index was associated with the estimated instantaneous reproductive number two weeks later (figure 3). A 10% lower mean mobility index was associated with a decrease in the instantaneous reproductive number of 0.061 2 weeks later (*β* = −0·061; 95% CI: −0·071, −0·052). When the model was adjusted for days since the 100^th^ case, the association persisted but was attenuated (*β* = −0·029; 95% CI: −0·044, −0·014). Using a 3-week lag for mobility produced a slightly weaker strength of association (table 1). Excluding the first week of case data (March 23) produced relatively comparable results (table S3).

**Figure 3.**
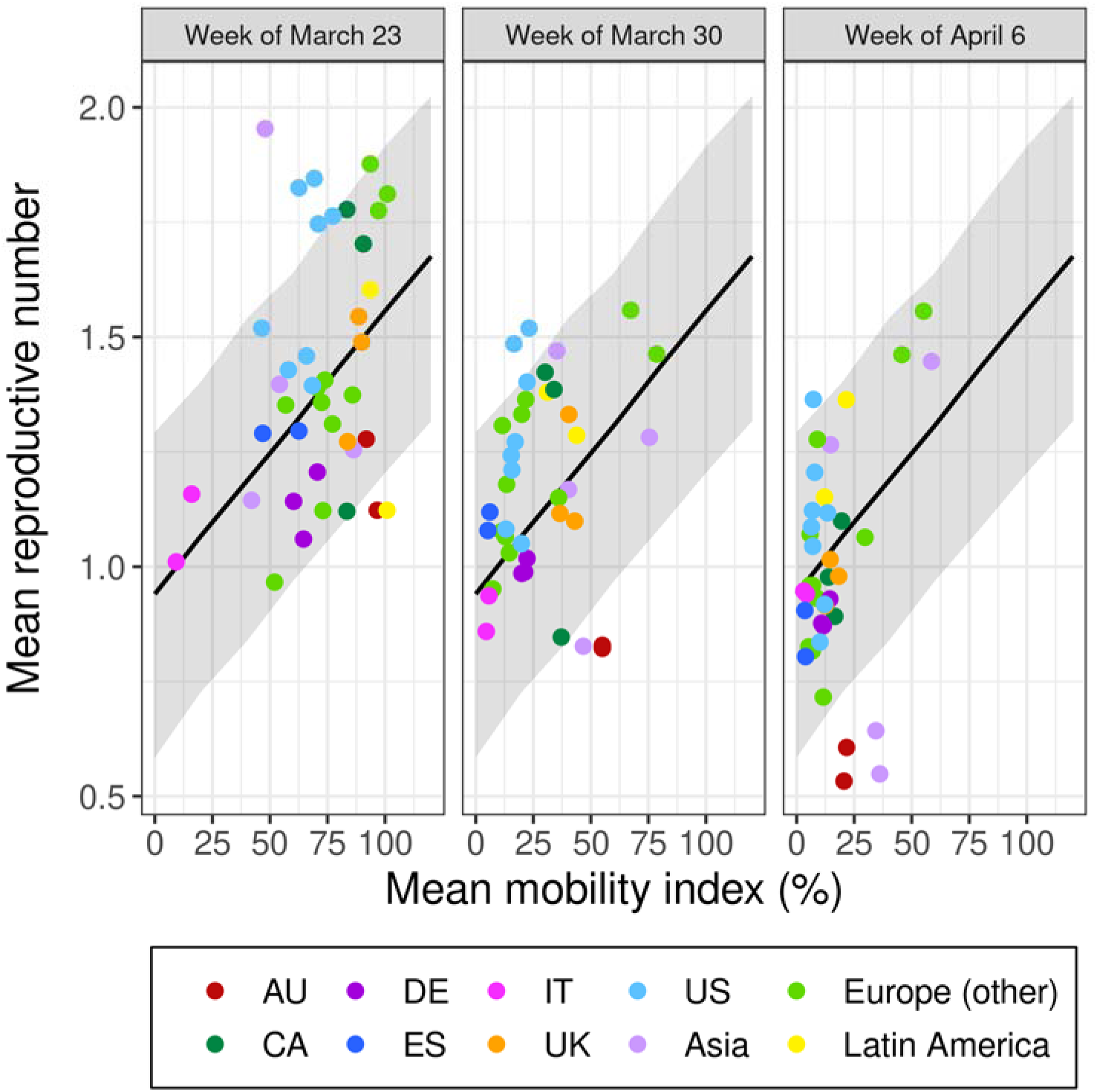
The association between the mean instantaneous reproductive number (aggregated by week) and mean mobility index two weeks prior for 40 states and countries. Each pair of weeks (labeled according to the week the reproductive number was estimated) is plotted separately for a model fit on all three weeks of data. The 95% prediction interval is shown in gray. AU = Australia; CA = Canada; DE = Germany; ES = Spain; IT = Italy; UK = United Kingdom; US = United States.

## Discussion

We found that a mobility index of public transit users in cities spanning 5 continents predicted growth in reported cases of COVID-19 2 to 3 weeks later. Such an index could be used by public health and governments attempting to understand the impacts of physical distancing and mobility restriction measures during the COVID-19 pandemic.

While the metric we evaluated is predictive, its availability is limited to a handful of cities located mainly in Europe and North America, although many of these cities are major regional epicenters of infection. The index also reflects the movement of a limited portion of the population—transit users—and provides no insight into the number and distribution of close contacts that could lead to transmission. Although we have justified the use of a city-level mobility metric above, this inevitably introduces measurement error for an outcome aggregated at the national or sub-national level. Variation across countries and regions in the delay between symptom onset and public reporting of cases adds uncertainty regarding the correct lag between changes in mobility and the resulting effects on growth rates. This should become less of an issue as more rapid and standardized testing is implemented across regions.

Our study had other limitations. Our analysis does not confirm a causal pathway through mobility, but rather a strong association that warrants further evaluation. For example, it is possible that countries successfully enforcing physical distancing are also more successfully implementing interventions such as contact tracing or widespread testing, which may also contribute to the observed association. We also did not account for imported cases in the calculation of the instantaneous reproductive number; however, locally acquired cases were certainly undercounted during this period, and likely to a greater degree than imported cases due to the increased attention on international travelers. Further, imported cases are expected to account for an increasingly small proportion of total cases in the latter two weeks (March 30 and April 6) due to the implementation of travel restrictions in mid–late March. Re-running the models to exclude the first week (March 23) of case data produced relatively similar results to the full models (for both growth rate and reproductive number), suggesting that imported cases do not drive our results.

Additional measures of human mobility and physical distancing are urgently needed in order to better understand the impacts of these policies on transmission dynamics. For example, Lasry and colleagues^18^ examined the relationship between the percentage of cell phone users leaving their homes and COVID-19 case trajectories in 4 metropolitan areas in the United States. Corporations such as Google and Apple have released global mobility data that may contain a greater variety of contact and mobility patterns, and research is ongoing to evaluate the usefulness of these metrics.^19–21^ These metrics need not necessarily be granular, as physical distancing measures are usually implemented at a broad scale. Further evaluation of the utility of these metrics in guiding population interventions are needed, particularly for supporting the steps necessary to keep the reproductive number of the disease below 1.

Though necessary, these strategies are already proving to have dire consequences on other aspects of health and well-being.^22–25^ The value of mobility metrics is set to increase dramatically as jurisdictions gradually re-open and consider alternative strategies such as intermittent physical distancing.^26^ Ideally, more targeted measures such as aggressive contact tracing, testing, and isolation programs will eventually decouple mobility from COVID-19 transmission.^3,27^ Until then, we hope our results will be helpful to reassure the public that, despite the immense economic, social and psychological costs, their continued cooperation with existing public health measures will have powerful long-term benefits.

## Data Availability

Data and code to reproduce the analysis are available at: https://github.com/jeanpaulrsoucy/covid-19-mobility. This study used exclusively publicly available data (sources listed in the supplementary material).

## Author contributions

KAB and J-PRS conceived of the study. J-PRS, SLS, IB, and DJW curated the data. J-PRS and KAB performed the analysis. J-PRS, SLS, KAB, and IB wrote the initial draft. All authors contributed to and approved the final manuscript.

## Dating sharing

Data and code to reproduce the analysis are available at: https://github.com/ieanpaulrsoucv/covid-19-mobilitv. This study used exclusively publicly available data (sources listed in the supplementary material).

## Supplementary Material

**Figure S1.**
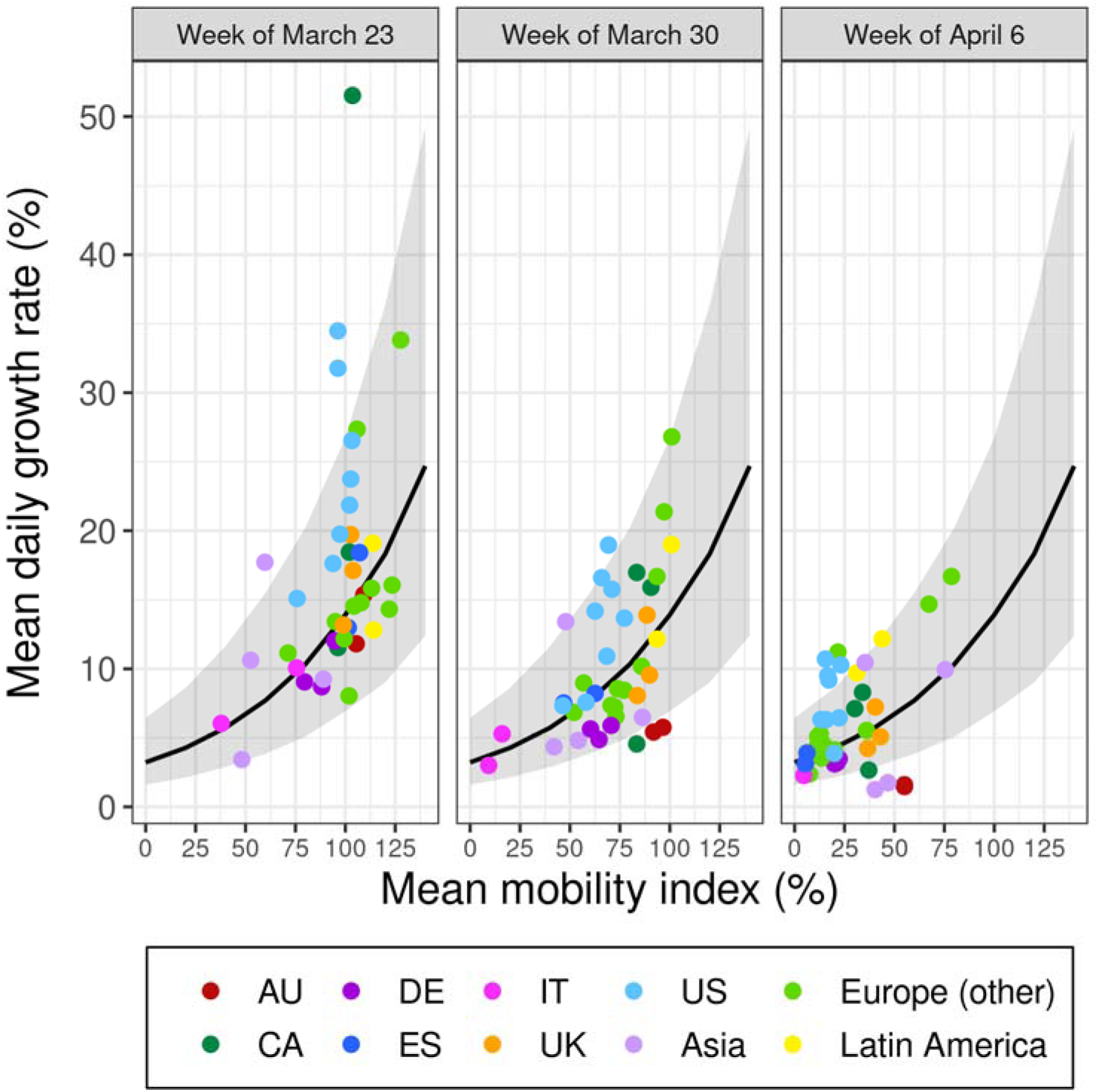
The association between mean daily growth rate (aggregated by week) and mean mobility index three weeks prior for 40 states and countries. Each pair of weeks (labeled according to the week growth rate was measured) is plotted separately for a model fit on all three weeks of data. The 95% prediction interval is shown in gray. AU = Australia; CA = Canada; DE = Germany; ES = Spain; IT = Italy; UK = United Kingdom; US = United States.

**Figure S2.**
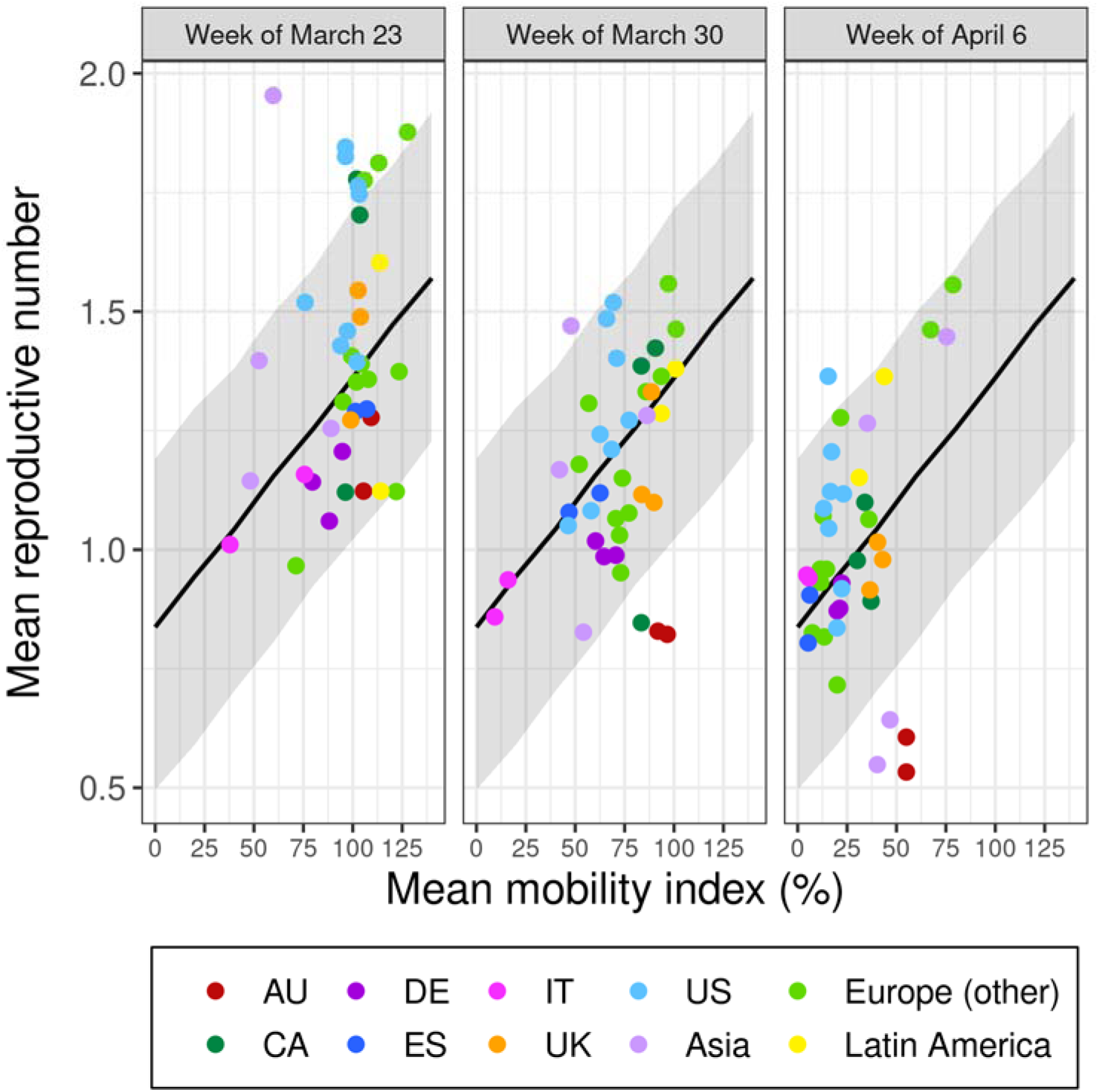
The association between the mean instantaneous reproductive number (aggregated by week) and mean mobility index three weeks prior for 40 states and countries. Each pair of weeks (labeled according to the week the reproductive number was estimated) is plotted separately for a model fit on all three weeks of data. The 95% prediction interval is shown in gray. AU = Australia; CA = Canada; DE = Germany; ES = Spain; IT = Italy; UK = United Kingdom; US = United States.

**Table S1.**
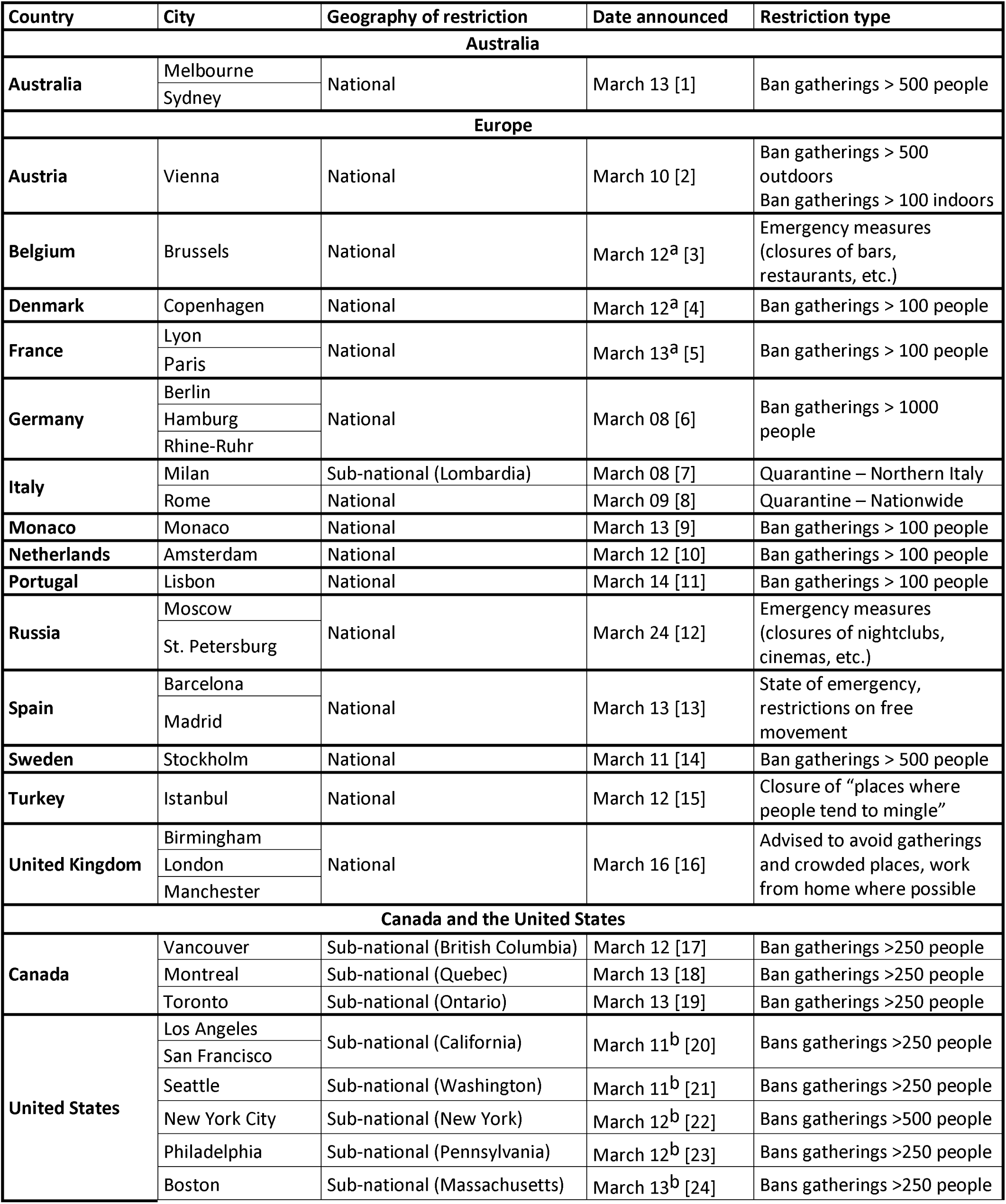
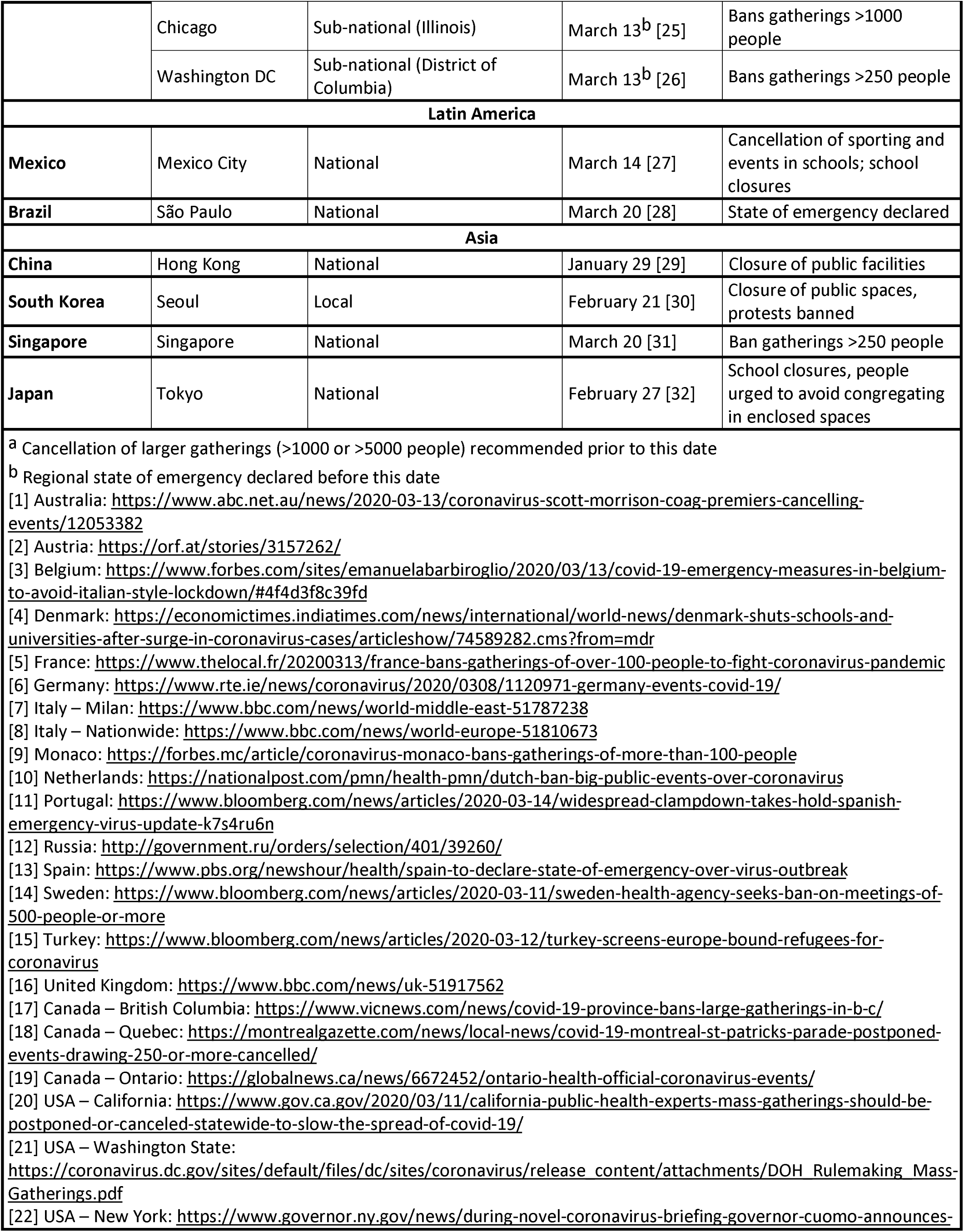
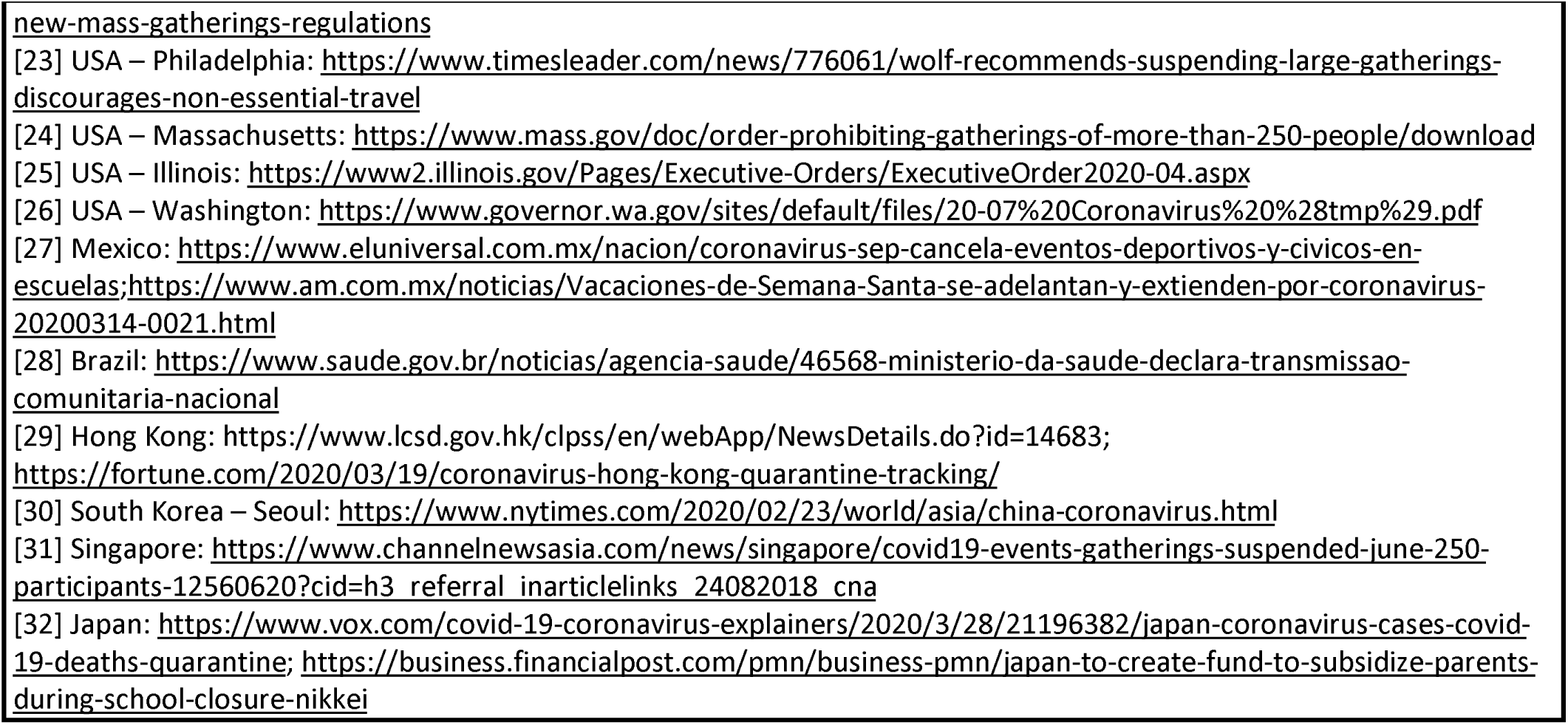
Date when the first major physical distancing measure (restrictions on public gatherings or mandatory business closures) was announced by state or country.

**Table S2.**
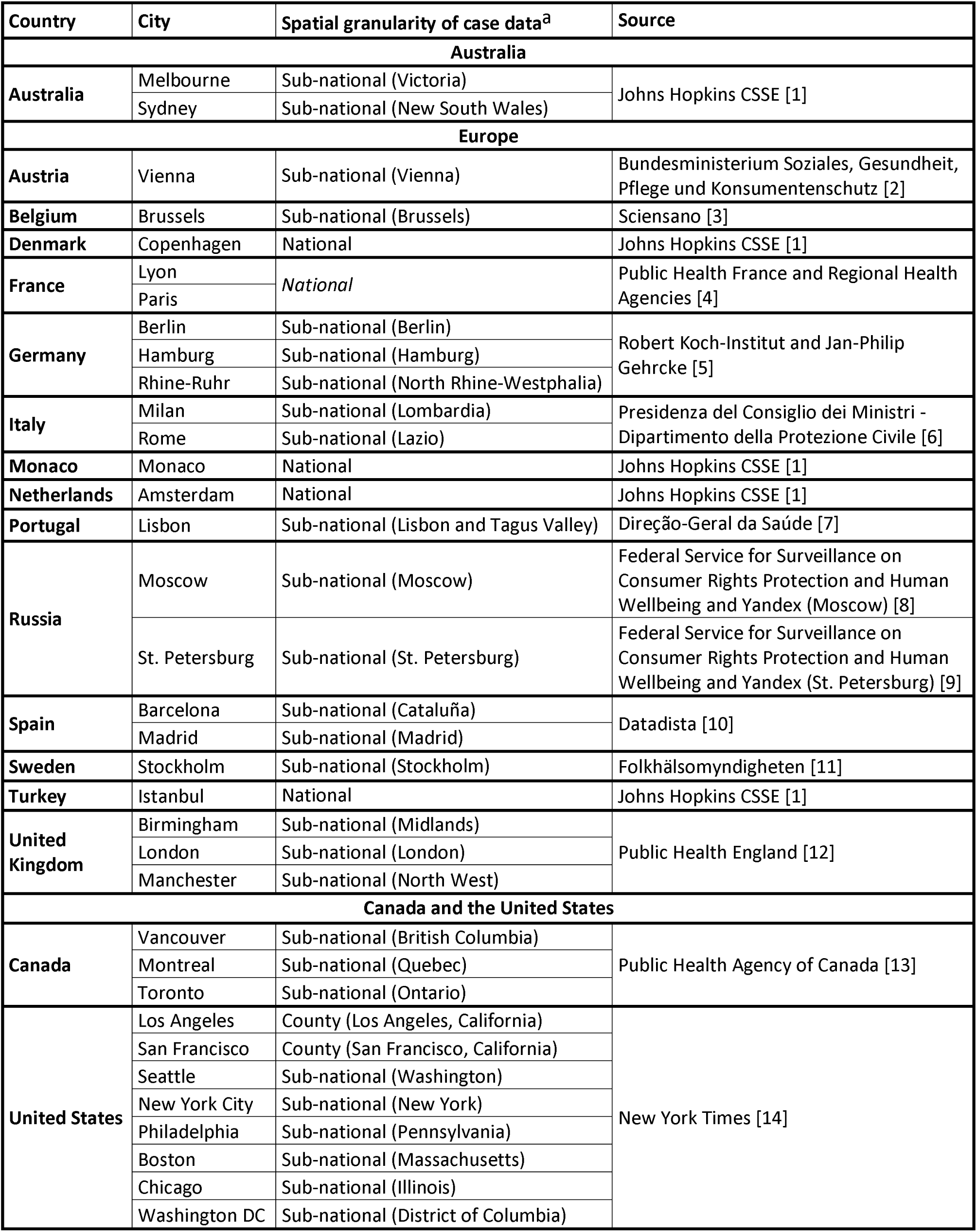
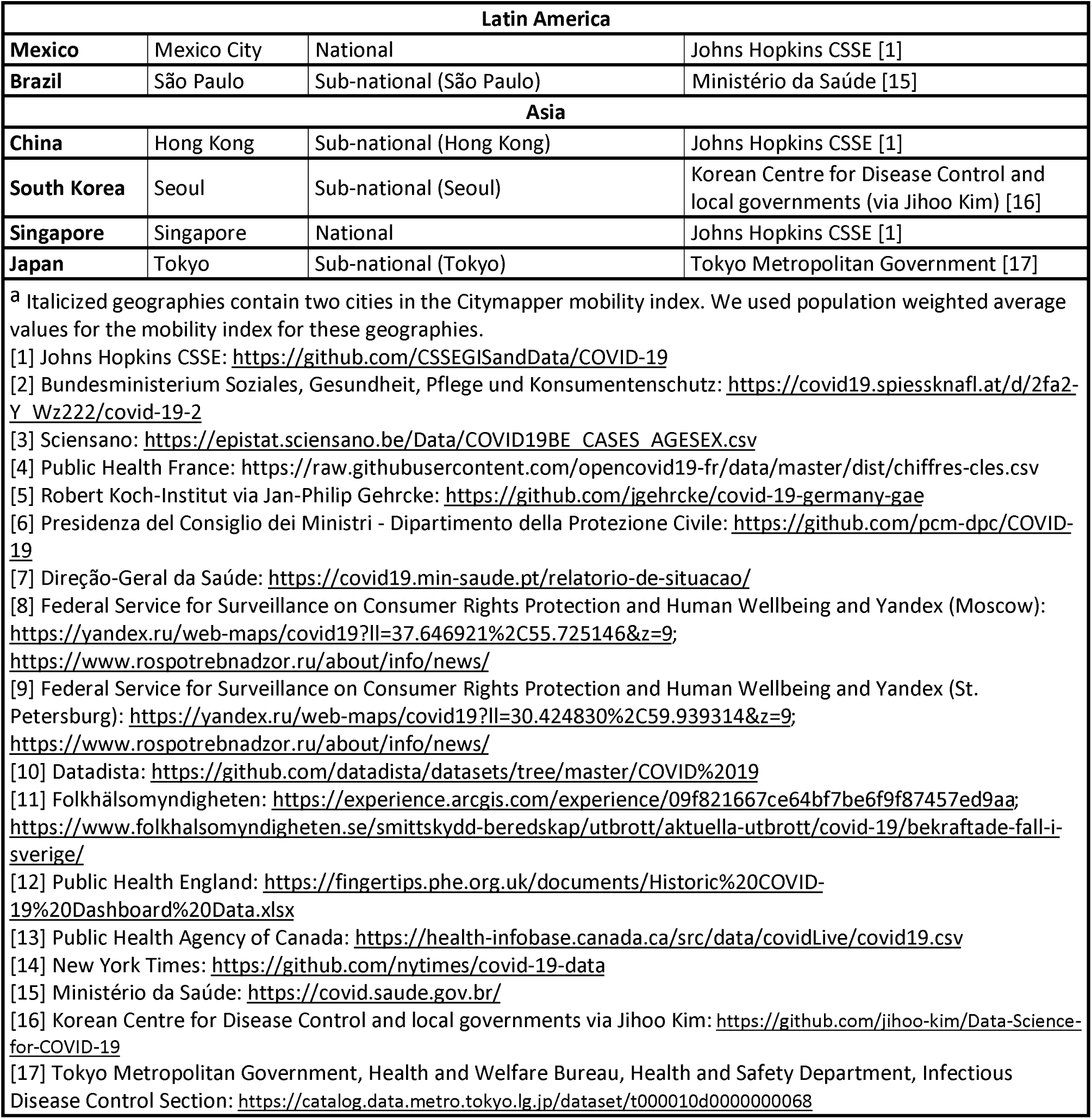
Sources for cumulative case data and whether they were obtained at a national or sub-national level.

| Country | City | Spatial granularity of case data<sup>a</sup> | Source |
| --- | --- | --- | --- |
| Australia |  |  |  |
| Australia | Melbourne | Sub-national (Victoria) | Johns Hopkins CSSE [1] |
|  | Sydney | Sub-national (New South Wales) |  |
| Europe |  |  |  |
| Austria | Vienna | Sub-national (Vienna) | Bundesministerium Soziales, Gesundheit, Pflege und Konsumentenschutz [2] |
| Belgium | Brussels | Sub-national (Brussels) | Sciensano [3] |
| Denmark | Copenhagen | National | Johns Hopkins CSSE [1] |
| France | Lyon | National | Public Health France and Regional Health Agencies [4] |
|  | Paris |  |  |
| Germany | Berlin | Sub-national (Berlin) | Robert Koch-Institut and Jan-Philip Gehrcke [5] |
|  | Hamburg | Sub-national (Hamburg) |  |
|  | Rhine-Ruhr | Sub-national (North Rhine-Westphalia) |  |
| Italy | Milan | Sub-national (Lombardia) | Presidenza del Consiglio dei Ministri - Dipartimento della Protezione Civile [6] |
|  | Rome | Sub-national (Lazio) |  |
| Monaco | Monaco | National | Johns Hopkins CSSE [1] |
| Netherlands | Amsterdam | National | Johns Hopkins CSSE [1] |
| Portugal | Lisbon | Sub-national (Lisbon and Tagus Valley) | Direção-Geral da Saúde [7] |
| Russia | Moscow | Sub-national (Moscow) | Federal Service for Surveillance on Consumer Rights Protection and Human Wellbeing and Yandex (Moscow) [8] |
|  | St. Petersburg | Sub-national (St. Petersburg) | Federal Service for Surveillance on Consumer Rights Protection and Human Wellbeing and Yandex (St. Petersburg) [9] |
| Spain | Barcelona | Sub-national (Cataluña) | Datadista [10] |
|  | Madrid | Sub-national (Madrid) |  |
| Sweden | Stockholm | Sub-national (Stockholm) | Folkhälsomyndigheten [11] |
| Turkey | Istanbul | National | Johns Hopkins CSSE [1] |
| United Kingdom | Birmingham | Sub-national (Midlands) | Public Health England [12] |
|  | London | Sub-national (London) |  |
|  | Manchester | Sub-national (North West) |  |
| Canada and the United States |  |  |  |
| Canada | Vancouver | Sub-national (British Columbia) | Public Health Agency of Canada [13] |
|  | Montreal | Sub-national (Quebec) |  |
|  | Toronto | Sub-national (Ontario) |  |
| United States | Los Angeles | County (Los Angeles, California) | New York Times [14] |
|  | San Francisco | County (San Francisco, California) |  |
|  | Seattle | Sub-national (Washington) |  |
|  | New York City | Sub-national (New York) |  |
|  | Philadelphia | Sub-national (Pennsylvania) |  |
|  | Boston | Sub-national (Massachusetts) |  |
|  | Chicago | Sub-national (Illinois) |  |
|  | Washington DC | Sub-national (District of Columbia) |  |

| Latin America |  |  |  |
| --- | --- | --- | --- |
| **Mexico** | Mexico City | National | Johns Hopkins CSSE [1] |
| **Brazil** | São Paulo | Sub-national (São Paulo) | Ministério da Saúde [15] |
| Asia |  |  |  |
| **China** | Hong Kong | Sub-national (Hong Kong) | Johns Hopkins CSSE [1] |
| **South Korea** | Seoul | Sub-national (Seoul) | Korean Centre for Disease Control and local governments (via Jihoo Kim) [16] |
| **Singapore** | Singapore | National | Johns Hopkins CSSE [1] |
| **Japan** | Tokyo | Sub-national (Tokyo) | Tokyo Metropolitan Government [17] |

<sup>a</sup> Italicized geographies contain two cities in the Citymapper mobility index. We used population weighted average values for the mobility index for these geographies.
[1] Johns Hopkins CSSE: https://github.com/CSSEGISandData/COVID-19
[2] Bundesministerium Soziales, Gesundheit, Pflege und Konsumentenschutz: https://covid19.spiessknafl.at/d/2fa2-Y_Wz222/covid-19-2
[3] Sciensano: https://epistat.sciensano.be/Data/COVID19BE_CASES_AGESEX.csv
[4] Public Health France: https://raw.githubusercontent.com/opencovid19-fr/data/master/dist/chiffres-cles.csv
[5] Robert Koch-Institut via Jan-Philip Gehrcke: https://github.com/jgehrcke/covid-19-germany-gae
[6] Presidenza del Consiglio dei Ministri - Dipartimento della Protezione Civile: https://github.com/pcm-dpc/COVID-19
[7] Direção-Geral da Saúde: https://covid19.min-saude.pt/relatorio-de-situacao/
[8] Federal Service for Surveillance on Consumer Rights Protection and Human Wellbeing and Yandex (Moscow): https://yandex.ru/web-maps/covid19?ll=37.646921%2C55.725146&z=9;https://www.rosпотребнадзор.ru/about/info/news/
[9] Federal Service for Surveillance on Consumer Rights Protection and Human Wellbeing and Yandex (St. Petersburg): https://yandex.ru/web-maps/covid19?ll=30.424830%2C59.939314&z=9;https://www.rosпотребнадзор.ru/about/info/news/

**Table S3.** Model coefficients (with 95% credible intervals) for the association between a 10% decrease in the mobility index and the mean daily growth rate or instantaneous reproductive number (both aggregated by week) assuming a lag of two or three weeks, using outcome data excluding the week of March 23. Model coefficients are presented with and without adjustment for days since 100^th^ case. Models include the second and third weeks of data (March 30–April 12) for 40 countries and states (39 in the adjusted models). CI = credible interval.

| Mean daily growth rate (%) ($\exp(\beta)$) | | | |
| --- | --- | --- | --- |
| Mobility index | Unadjusted (95% CI) | Adjusted (95% CI)<sup>a</sup> |  |
|  | Mobility<sup>b</sup> | Mobility<sup>b</sup> | Days since 100 cases<sup>c</sup> |
| 2-week lag | 0.780 (0.722, 0.845) | 0.913 (0.852, 0.980) | 0.554 (0.478, 0.644) |
| 3-week lag | 0.884 (0.865, 0.903) | 0.938 (0.908, 0.971) | 0.668 (0.551, 0.800) |
| Instantaneous reproductive number ($\beta$) | | | |
| Mobility index | Unadjusted (95% CI) | Adjusted (95% CI)<sup>a</sup> |  |
|  | Mobility<sup>b</sup> | Mobility<sup>b</sup> | Days since 100 cases<sup>c</sup> |
| 2-week lag | -0.065 (-0.093, -0.037) | -0.039 (-0.068, -0.010) | -0.137 (-0.195, -0.079) |
| 3-week lag | -0.038 (-0.049, -0.028) | -0.026 (-0.040, -0.011) | -0.084 (-0.154, -0.015) |

<sup>a</sup> Excludes the Principality of Monaco.
<sup>b</sup> Coefficient for a 10% decrease in the mobility index.
<sup>c</sup> Coefficient for a 10-day increase since the 100<sup>th</sup> reported case.

